# Exploring interaction between genetically predicted body mass index and serum 25-hydroxyvitamin D levels on the odds for psoriasis in UK Biobank and the HUNT Study: A factorial Mendelian randomisation study

**DOI:** 10.1101/2024.07.01.24309489

**Authors:** Marita Jenssen, Nikhil Arora, Mari Løset, Bjørn Olav Åsvold, Laurent Thomas, Ole-Jørgen Gangsø Bekkevold, Xiao-Mei Mai, Yi-Qian Sun, Anne-Sofie Furberg, Rolf Jorde, Tom Wilsgaard, Kjersti Danielsen, Ben Michael Brumpton

**Author notes:** These authors share the last authorship. Corresponding authors: Ben Michael Brumpton, Marita Jenssen.

## Abstract

**Background:** Mendelian randomisation (MR) studies show that higher body mass index (BMI) and lower 25-hydroxyvitamin D (25[OH]D) increase psoriasis risk. The combined effect of these factors has not been explored using factorial MR.

**Methods:** Using cross-sectional data from UK Biobank (UKB, n=398 404) and the Trøndelag Health Study (HUNT, n=86 648), we calculated polygenic risk scores for BMI and 25(OH)D to estimate odds ratios for psoriasis using 2×2 and continuous factorial MR. We quantified additive interaction by relative excess risk due to interaction (RERI)-estimates. We also performed traditional observational analyses in UKB.

**Results:** There were 12 207 (3.1%) participants with psoriasis in UKB and 7794 (9.0%) in HUNT. In 2×2 factorial MR, we found no evidence of relative excess risk for psoriasis due to interaction between genetically predicted higher BMI and lower 25(OH)D, neither in UKB (RERI −0.01, 95% confidence interval (CI) −0.08, 0.07) nor in HUNT (RERI −0.04, 95% CI −0.14, 0.06). The same was observed in the continuous factorial MR and observational analyses.

**Conclusions:** This study did not find evidence of interaction between BMI and 25(OH)D on the risk of psoriasis. Given minor differences in measured BMI and 25(OH)D between groups, small effects may have been undetected.

## Introduction

Psoriasis is a chronic, inflammatory skin disease which is associated with multiple comorbidities and has considerable impact on quality of life^1^. The prevalence varies between populations and geographical locations, and is estimated to be 2-4% among adults in Western countries^2^. However, life-time prevalence as high as 6.6-11.4% has been reported in Norway^3,4^. Some data indicate increasing prevalence over the past decades^4-6^.

The obesity epidemic may partly explain the rising psoriasis prevalence, as studies have consistently demonstrated an association between obesity and increased risk of psoriasis^7-9^. Previous Mendelian randomisation (MR) studies, including European and Japanese populations, have demonstrated a causal relationship from higher body mass index (BMI) to an increased risk of psoriasis^10,11^. Where reverse causality was found to be unlikely^11^.

Vitamin D has several effects of importance to skin physiology; of particular relevance to psoriasis is regulation of the immune system, as well as proliferation and maturation of keratinocytes^12^. These physiological effects are utilized when treating psoriasis with topical vitamin D analogues^1^. Observational studies has found an association between lower serum 25-hydroxyvitamin D (25[OH]D) and psoriasis^13^. MR studies have additionally demonstrated a causal relationship from lower 25(OH)D to an increased risk of psoriasis^14,15^. Moreover, no reverse causal link has been observed^16^.

Recent observational studies suggest that the association between 25(OH)D and psoriasis may be modified by adiposity (BMI and hip-waist ratio)^14,17,18^. No previous studies have explored this possible combined effect (i.e. interaction) using MR designs, which are considered more robust to residual confounding and reverse causality than traditional observational approaches^19^. In factorial MR, genetic variants are used as instrumental variables to assess interaction between modifiable risk factors^20,21^. Understanding relative excess (disease) risk due to interaction (RERI) can have implications for targeted interventions and public health strategies^22^.

In this study, we investigated RERI (hereafter referred to as interaction) between genetically predicted BMI and 25(OH)D on psoriasis using observational data from two independent population-based cohorts; The UK Biobank (UKB) and the Trøndelag Health Study (HUNT). We also explored interaction between measured 25(OH)D and BMI on the odds of psoriasis using a traditional cross-sectional design in UKB.

## Materials and methods

### UK Biobank

UKB is a prospective cohort study including data from approximately 500 000 adult UK citizens (40-69 years old at recruitment) enrolled between March 2006 and July 2010^23^. Participants (5.5% of invitees) completed a touchscreen questionnaire, a formal interview, standardised clinical examinations, and biological sampling. Genotyping was performed, and data linked with health records, including hospital records, death registry, and general practitioner data. The latter was available for 45% of participants when we accessed data in May 2023. Details regarding data collection has been described previously^24^. UKB data-fieldIDs used in this study are listed in the Online supplementary.

We restricted the UKB sample to include white British and unrelated individuals (kinship coefficient cut-off 0.0884) with complete data on the relevant SNPs and valid psoriasis code (flow-chart in supplementary Figure S1). In total 398 404 participants (214 178 women and 184 226 men) were included in the analysis.

### The HUNT Study

HUNT is a population-based cohort study to which adults (≥20 years old at recruitment) in Trøndelag County, Norway are invited to repeated surveys^25,26^. Data is collected through questionnaires, interviews, standardized clinical examinations, biological sampling, and linkage with health records. The number of participants (attendance proportion) was 77 202 (89.4%) in HUNT1 (1984–1986), 65 228 (69.5%) in HUNT2 (1995–1997), 50 800 (54.1%) in HUNT3 (2006–2008), and 56 042 (54.0%) in HUNT4 (2017–2019)^25^. Among the participants in HUNT2-4, 88 615 individuals have been genotyped^27^. We excluded non-European participants, those with invalid psoriasis code or missing information on age (flow-chart in supplementary Figure S2). In total 86 648 participants (45 949 women and 40 699 men) were included in the analysis.

### Classification of psoriasis

Participants with reported psoriasis either in medical records (ICD codes and/or codes from primary care) or by self-report were classified as having psoriasis.

In UKB we used Date L40 first reported (psoriasis) which combines information from death records, hospital records, primary care data and self-report. The latter was registered if the participant mentioned psoriasis during the formal interview when asked about doctor diagnosed serious illnesses or disabilities.

In HUNT, ICD-codes (ICD9: 696.0/1, ICD10: L40) were available from hospitals and private specialist practitioners and ICPC-2 codes (S91) from general practitioners. Self-reported psoriasis was defined by answering “yes” to the questionnaire question “Have you had, or do you have psoriasis?”. Self-reported psoriasis has acceptable validity in HUNT (positive predictive value 78%; 95% confidence interval [CI] 69–85)^28^. The source psoriasis diagnosis for the included participants are listed in supplementary table S1 and figure S3.

### Genetic instruments

In UKB, participants were genotyped using one of two genotyping arrays; The Applied Biosystems UK BiLEVE Axiom Array or Applied Biosystems UK Biobank Axiom Array. In HUNT, genotyping was performed using one of four Illumina HumanCoreExome arrays; HumanCoreExome12 version 1.0, HumanCoreExome12 version 1.1, UM HUNT Biobank version 1.0 or UM HUNT Biobank v2.0. The quality control and the imputation procedures have been described in detail previously^27,29,30^.

Polygenic risk scores (PRSs) summarise genetic information from several variants into one single variable, with the advantage of explaining a larger amount of variation in a trait compared to single variants^31^. We calculated PRSs for BMI and 25(OH)D (hereafter called BMI-PRS and vitD-PRS, respectively) for each participant as the sum of trait increasing alleles, weighted by the GWAS-effect size of the respective SNP^31^. For our primary analysis, we used a BMI-PRS combining 941 SNPs^32^. In sensitivity analysis, we used another BMI-PRS combining 77 SNPs^33^. For the vitD-PRS we presented combinations of SNPs used in previous MR studies^34-36^. In our primary analysis the vitD-PRS included 21 SNPs from genes with well-known functions in the vitamin D metabolism (CYP2R1, DHCD7, CYP24A1 and GC)^37^. In sensitivity analyses, we used vitD-PRSs combining 35 SNPs^34,35^, and 71 SNPs^37^, respectively. A more detailed description of our selection of SNPs is found in the Online supplementary. Both BMI-PRSs and the vitD-PRS used in our primary analyses have previously been used in one-sample MR studies with psoriasis as outcome, and found to be robust to pleiotropy^11,37^.

### Other measurements

Age was defined by age at first inclusion.

BMI was calculated as weight in kilograms per height in meters squared. Standing height and weight were measured at the assessment centres. For participants with repeated measurements, the first measurement was used. BMI was dichotomised using WHO’s proposed threshold for public health action (BMI >27.5 kg/m^2^)^38^.

Serum 25(OH)D was measured in UKB using a certified chemiluminescence immunoassay (DiaSorin LIAISON XL, Italy)^39^. Non-fasting blood samples collected at the assessment centres were stored at minus 80oC before analysis. Season of blood draw was used for seasonal adjustment (December to February = Winter; March to May = Spring; June to August = Summer; September to November = Autumn). All measurements were converted to autumn-measurement by subtracting the respective season mean and adding the autumn mean (e.g., a summer value of 80 nmol/L was converted to season adjusted value by this formula [80 – summer mean + autumn mean])^15,37^. In HUNT, 25(OH)D measurements were only available for a limited number of participants^40^, and therefore not included in the analysis.

### Statistical analyses

Logistic regression was used to estimate odds ratios (ORs) for psoriasis using individual-level data separately in UKB and HUNT.

We dichotomized the BMI-PRS and vitD-PRS by the respective median value (separately for UKB and HUNT). Values equal to or below the median represented low PRS and values above median represented high PRS. Thereafter, participants were separated in four groups based on the dichotomised BMI-PRS and vitD-PRS categories (Figure 1). We expected those with low BMI-PRS and high vitD-PRS (representing lower BMI and higher 25[OH]D) to have lowest odds for psoriasis and chose this group as reference.

**Figure 1:**
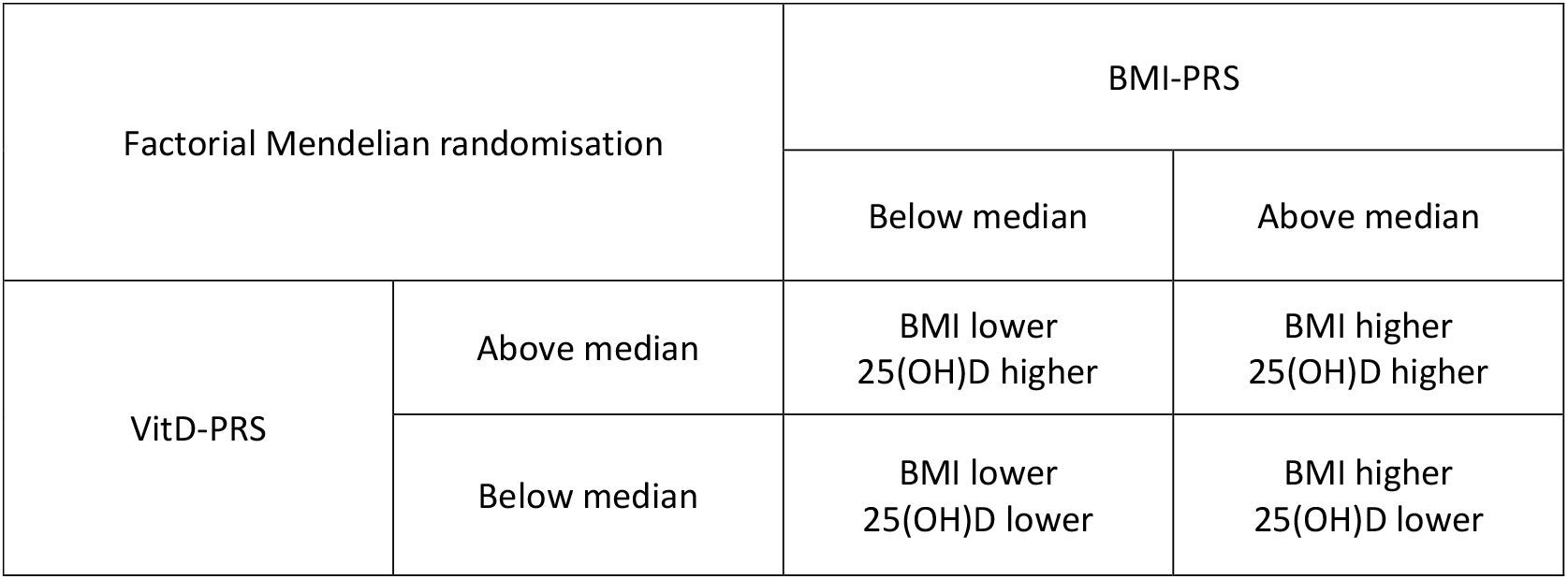
Factorial Mendelian randomisation using a 2×2 design. Adapted from *Rees JMB, Foley CN, Burgess S. Factorial Mendelian randomization: using genetic variants to assess interactions. International Journal of Epidemiology 2019; 49: 1147-58117^20^*. BMI=body mass index. PRS=polygenic risk score. 25(OH)D=25-hydroxyvitamin D. BMI-PRS = the PRS for BMI. VitD-PRS = the PRS for 25(OH)D.

We performed 2×2 factorial MR analyses using the four groups based on the dichotomised BMI-PRS and vitD-PRS as an exposure variable. We also performed continuous factorial MR including BMI-PRS, vitD-PRS (both standardised to have a mean of 0 and standard deviation of 1) and their cross-product as predictors. All models included age, sex, genetic batch and 20 principal components^27,30^ (to account for population stratification). To ease comparison with measured BMI and 25(OH)D, we also present the OR estimates (UKB) scaled to represent 5 kg/m^2^ increase in BMI and 10 nmol/L decrease in 25(OH)D.

In UKB, we also performed cross-sectional analyses using observational data. In the continuous approach, predictors were measured 25[OH]D and BMI, and their cross-product. In the 2×2 approach, the predictor was four groups based on dichotomised 25(OH)D (≥ or <25 nmol/L) and BMI (> or ≤27.5 kg/m^2^), using 25(OH)D ≥25 nmol/L + BMI ≤27.5 kg/m^2^ as a reference group. Adjusted models included age and sex.

Interaction on an additive scale was assessed by estimating RERI^41,42^. A RERI estimate of zero supports exact additivity, above zero supports positive interaction, and below zero supports negative interaction.

Univariable linear models were applied to assess the association between the respective PRSs and measured 25(OH)D (UKB) and BMI (UKB and HUNT). We evaluated F-statistics and R^2^ estimates to assess the strength of the genetic instruments. Moreover, we explored the association between the PRSs and possible confounders (BMI, age, sex) to assess the second MR assumption (i.e. the instrument is not associated with confounders). Additionally, the factorial MR was repeated with multiple PRSs to explore potential pleiotropy.

PLINK 2.0 was used to generate PRSs in UKB^43^. Generation of PRSs in HUNT, and all statistical analyses were performed in R version 4.2.1 (R Foundation for Statistical Computing, Vienna, Austria). Estimates are reported with a 95% CI.

### Ethics

All participants have provided written informed consent. The National Health Service National Research Ethics Service approved the UK Biobank (reference 11/NW/0382). The Regional Committee for Medical and Health Research Ethics (REK) Central approved the HUNT Study. The present study was conducted using the UKB Resource under application number 40135, and was approved by the REK, Mid-Norway (2015/2003).

## Results

### General characteristics

The UKB sample included 398 404 individuals (53.8% women) with mean (SD) age 56.9 (7.9) years, BMI 27.4 (4.8) kg/m^2^, and season adjusted 25(OH)D 54.8 (19.7) nmol/L (Table 1). The HUNT sample included 86 648 individuals (53.0% women) with mean (SD) age 46.1 (16.9) years and BMI 26.4 (4.4) kg/m^2^ (Table 1).

**TABLE 1.** Characteristics of the participants; UK Biobank (n=398 404^*^) and HUNT (n= 86 648)

|  | UK Biobank |  | HUNT |  |
| --- | --- | --- | --- | --- |
|  | No psoriasis | Psoriasis<sup>1</sup> | No psoriasis | Psoriasis<sup>1</sup> |
| Number of participants | 386 197 / 96.9 | 12 207 / 3.1 | 78 854 / 91.0 | 7 794 / 9.0 |
| Age (years) | 56.9 (7.9) | 57.2 (7.8) | 46.2 (17.1) | 44.7 (13.8) |
| Sex |  |  |  |  |
| Female | 208 122 / 53.9 | 6 056 / 49.6 | 41 674 / 52.8 | 4 275 / 54.8 |
| Male | 178 075 / 46.1 | 6 151 / 50.4 | 37 180 / 47.2 | 3 519 / 45.2 |
| 25(OH)D (nmol/L)<sup>2</sup> | 54.8 (19.7) | 53.3 (19.7) | NA | NA |
| BMI (kg/m<sup>2</sup>) | 27.4 (4.7) | 28.3 (5.1) | 26.4 (4.3) | 27.0 (4.6) |

Continuous variables given as mean (SD) and categorical as number / proportions.
HUNT = The Trøndelag Health Study. 25(OH)D = serum 25-hydroxyvitamin D. BMI = body mass index. NA=not applicable.
<sup>1</sup> Psoriasis = Medical records and/or self-report
<sup>2</sup> Season-adjusted (Autumn mean)
\*Dataset restricted (omitted participants missing genetic data + unrelated).

The prevalence of psoriasis was 3.1% (n=12 207) in UKB and 9.0% (n=7794) in HUNT (Table 1). Participants with psoriasis had on average higher BMI (in both cohorts) and lower 25(OH)D (UKB only) compared to participants without psoriasis.

Table 2 displays general characteristics across the 2×2 factorial MR groups. Having a genetically predicted higher BMI and lower 25(OH)D combined corresponded to an average 1.61 kg/m^2^ higher BMI and 7.39 nmol/L lower 25(OH)D in UKB, and 1.28 kg/m^2^ higher BMI in HUNT (Figure 2). A 1 SD increase in the BMI-PRS corresponded to a 1.05 and 0.81 kg/m^2^ increase in measured BMI in UKB and HUNT, respectively. A 1 SD decrease in the vitD-PRS corresponded to a 4.22 nmol/L lower measured 25(OH)D in UKB.

**TABLE 2.** Basic characteristics of the participants across the 2×2 factorial genetic risk groups. The UK Biobank (n=398 404) and HUNT (n=86 648)

| Factorial Group | 0<br>BMI-PRS<sup>1</sup> <<br>median<br>VitD-prs<sup>2</sup> ><br>median | 1<br>BMI-PRS<sup>1</sup> <<br>median<br>VitD-prs<sup>2</sup> <<br>median | 2<br>BMI-PRS<sup>1</sup> ><br>median<br>VitD-prs<sup>2</sup> ><br>median | 3<br>BMI-PRS<sup>1</sup> ><br>median<br>VitD-prs<sup>2</sup> <<br>median |
| --- | --- | --- | --- | --- |
| **UKB** |  |  |  |  |
| Number of participants | 99 587 | 99 616 | 99 615 | 99 586 |
| Psoriasis | 2 895 / 2.9 | 2 983 / 3.0 | 3 101 / 3.1 | 3 228 / 3.2 |
| Age | 57.0 (7.9) | 56.9 (7.9) | 56.9 (7.9) | 56.9 (7.9) |
| Sex |  |  |  |  |
| Female | 53 650 / 53.9 | 53 757 / 54.0 | 53 266 / 53.5 | 53 505 / 53.7 |
| Male | 45 937 / 46.1 | 46 859 / 46.0 | 46 349 / 46.5 | 46 081 / 46.3 |
| BMI<sup>3</sup> | 26.6 (4.5) | 26.9 (4.5) | 28.3 (5.0) | 28.2 (5.0) |
| 25(OH)D<sup>4</sup> | 58.5 (20.9) | 51.8 (17.8) | 57.6 (20.9) | 51.1 (17.8) |
| **HUNT** |  |  |  |  |
| Number of participants | 21 666 | 21 658 | 21 658 | 21 666 |
| Psoriasis | 1889 / 8.7 | 1875 / 8.7 | 2053 / 9.5 | 1977 / 9.1 |
| Age | 46.2 (17.0) | 46.2 (17.0) | 46.0 (16.8) | 45.9 (16.8) |
| Sex |  |  |  |  |
| Female | 11279 / 52.1 | 11583 / 53.5 | 11586 / 53.5 | 11501 / 53.1 |
| Male | 10387 / 47.9 | 10075 / 46.5 | 10072 / 46.5 | 10165 / 46.9 |
| BMI<sup>5</sup> | 25.8 (4.0) | 25.8 (4.1) | 27.1 (4.5) | 27.1 (4.6) |

<sup>1</sup> BMI-PRS=polygenic risk score for BMI, 940 SNPs in UKB/941 SNPs in HUNT
<sup>2</sup> VitD-prs=polygenic risk score for 25(OH)D, 21 SNPs in UKB/19 SNPs in HUNT
<sup>3</sup> Missing value in 1262
<sup>4</sup> Missing value in 35 389
<sup>5</sup> Missing value in 638
UKB=UK Biobank. HUNT=The Trøndelag Health Study.
Psoriasis = Medical records and/or self-reported.
BMI= body mass index (kg/m<sup>2</sup>). 25(OH)D = serum 25-hydroxyvitamin D (nmol/L), adjusted for season. SNP=single nucleotide polymorphism.

### Genetic instruments

The BMI-PRS (941 SNPs) explained 4.8% of the variation in BMI in UKB (R^2^ =4.8%, F-statistic=20180) and 3.4% of the variation in BMI in HUNT (R^2^ =3.4%, F-statistic=3065). It was weakly associated with 25(OH)D, age and sex in the UKB, and with age in HUNT (Supplementary table S2a).

The vitD-PRS (21 SNPs) explained 4.6% of the variation in 25(OH)D in UKB (R^2^ =4.6%, F-statistic=17490). It was weakly associated with BMI in the UKB, but not in HUNT (Supplementary table S2b).

### Factorial MR analysis

In 2×2 factorial MR analyses there was no evidence of relative excess risk for psoriasis due to interaction, neither in UKB (RERI[95% CI]=−0.01[−0.06, 0.09]) nor in HUNT (RERI[95% CI]=−0.04[−0.14, 0.06]) (Figure 2, supplementary table S3a and S3b).

**Figure 2.**
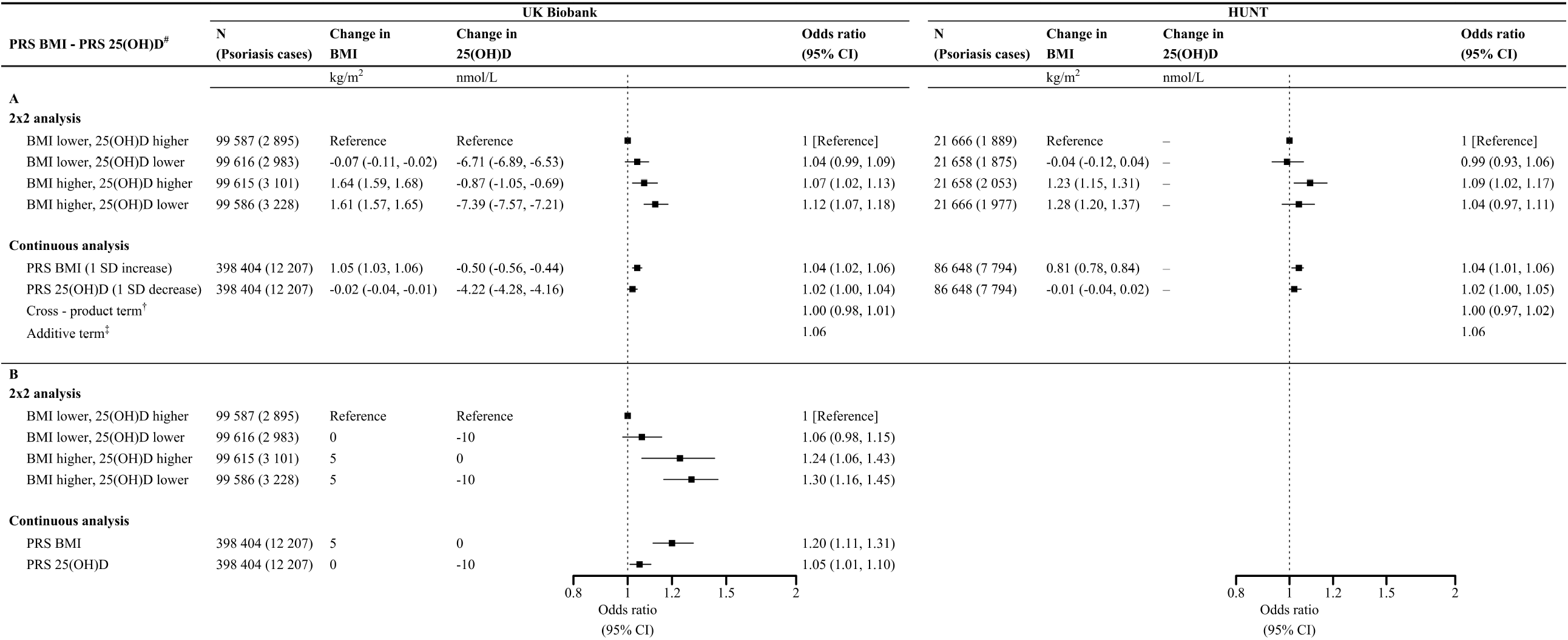
Odds ratio estimates for psoriasis from factorial Mendelian randomisation (MR) analyses in UK Biobank and HUNT. (A) Unscaled associations. (B) Associations in UKB scaled to represent a 5 kg/m2 increase in body mass index (BMI) and 10 nmol/L decrease in 25-hydroxyvitamin D (25[OH]D). Upper panel (A and B): Results from 2×2 factorial MR. Participants are separated in four groups based on median PRS for BMI and 25(OH)D^#^. Relative excess risk due to interaction (RERI, 95% CI) was estimated to −0.01(−0.06, 0.09) in UKB and −0.04(−0.14, 0.06) in HUNT. Interaction exists if RERI ≠ 0. Lower panel (A and B): Results from continuous factorial MR including the two PRSs on a continuous scale as well as the cross-product between PRSs. RERI (95% CI) was estimated to 0.00(−0.02, 0.02) in UKB and 0.00(−0.03, 0.02) in HUNT. Footnote: ^#^PRSs for body mass index and 25-hydroxyvitamin D. In 2×2 analysis PRSs were dichotomized at the median value, and combined to four groups. Odds ratio estimates derived from logistic regression models adjusted for age, sex, genetic batch and principal components 1 to 20. ^†^ Continuous factorial MR included the PRS for BMI, PRS for 25(OH)D (both standardised to have a mean of 0 and standard deviation of 1) and their cross-product. ^‡^ OR for the additive term of the continuous variables were calculated by exponentiating the sum of estimated beta-values for PRS for BMI (b1), PRS for 25(OH)D (b2) and their cross-product (b3) (i.e. OR = exp[b1+b2+b3]). HUNT = the Trøndelag Health Study. N=total number of participants. PRS=polygenic risk score. CI=confidence interval. SD=standard deviation.

Similarly, we observed no relative excess risk for psoriasis due to interaction in continuous factorial MR analysis assessing the combined effect of 1 SD increase in BMI-PRS (i.e., genetic risk for higher BMI), 1 SD decrease in vitD-PRS (i.e., genetic risk for lower 25[OH]D), and their cross-product, neither in UKB (RERI[95% CI]=0.00[−0.02, 0.02]), nor in HUNT (RERI[95% CI]=0.00[−0.03, 0.02]) (Figure 2 Supplementary table S4a and S4b).

In sensitivity analysis, no substantial differences in the RERI estimates were apparent across PRS-combinations in the 2×2 approach (Supplementary table S3a and S3b). In the continuous approach, RERI estimates were close to identical across PRS-combinations (Supplementary table S4a and S4b).

### Traditional observational analysis

In UKB, the combined impact of 10 nmol/L decrease in measured 25(OH)D, 5 kg/m^2^ increase in measured BMI, and their cross-product summed to 22% higher odds for psoriasis (OR[95% CI]=1.22[1.09, 1.37]), supporting exact additivity in the observational association (RERI[95% CI]=0.00[−0.01, 0.03]) (Supplementary table S5a). A similar trend was found when scaling the factorial MR analysis (Figure 2).

Support for exact additivity was also found in 2×2 observational analysis; having both 25(OH)D<25 nmol/L and BMI>27.5 was associated with 59% higher odds for psoriasis (OR[95% CI]=1.59[1.40, 1.80]) compared with having both 25(OH)D≥25 nmol/L and BMI≤27.5 (RERI[95% CI]=0.05[−0.21, 0.32]) (Supplementary table S5b).

## Discussion

### Main findings

In this factorial MR study, we found no evidence of relative excess risk for psoriasis due to interaction between genetically predicted higher BMI and lower 25(OH)D, neither in UKB nor in HUNT. Moreover, there was no evidence of relative excess risk for psoriasis due to interaction between measured BMI and 25(OH)D in the observational data from UKB. To our knowledge, no previous MR studies have investigated the interaction between BMI and 25(OH)D.

Previous MR studies investigating each factor separately have demonstrated a causal relationship between higher BMI^44^ and lower 25(OH)D^14,15^ and increased risk for psoriasis. These findings imply that strategies to reduce overweight and vitamin D deficiency in the population may influence the incidence of psoriasis. Complementing these previous findings, our results suggest that targeting both risk factors may be beneficial, but benefit cannot be expected to exceed the additive effect of the two. Although it is difficult to approximate the effect of interventions from MR estimates ^45-47^, they are valuable for testing causal associations and supporting effect direction of long-term interventions^46^. Public health recommendations for nutrition, physical activity and vitamin D supplementation are relevant examples of such life-long “interventions”, which would be extremely difficult to evaluate in trials (which might need follow-up for several decades). A North-American RCT showed a small preventive effect of vitamin D supplements for 5 years on autoimmune disease in general^48^. It was, however, underpowered to provide evidence for prevention of psoriasis specifically, as there were only 38 confirmed psoriasis cases among the ∼25 000 participants^48^. Longitudinal studies suggest that weight loss following bariatric surgery can prevent psoriasis in overweight subjects^49^. To our knowledge, no study has evaluated whether other weight reduction strategies have similar preventive effect.

Our findings do not support the effect modification between BMI and 25(OH)D suggested in previous observational studies^14,17^. However, direct comparison must be done cautiously, as inconsistencies may be explained by methodological differences; MR studies estimate the effect of small differences in life-long exposures, which may be incomparable to the impact of larger exposure differences evaluated over limited time-periods in traditional designs. Traditional observational designs can be affected by bias due to confounding, reverse causation, and measurement error, and violated or weak assumptions may bias MR estimates (discussed further below). That said, we found no substantial differences between the RERI estimates from MR and traditional analyses in our study, supporting the robustness of our findings. We do, however, recognize that potential interaction effects may be short-term, or manifested only at thresholds or extremes, such as very low 25(OH)D levels or in higher BMI categories. Moreover, interactions may only be relevant for disease activity in established psoriasis, as suggested by our recent study^50^, and not for incident psoriasis.

We have previously argued that overweight and vitamin D deficiency may act synergistically in driving inflammation in psoriasis, considering the link between obesity-associated low-grade inflammation and psoriasis, and the possible role of vitamin D deficiency in the pro-inflammatory state in obesity^7,50-52^. However, this hypothesis is not supported by the present findings.

### Strengths and limitations

The validity of MR effect estimates depend on three core assumptions; (1) Relevance; the instrument is robustly associated with the risk factor (exposure), (2) Independence; there is no association between the instrument and confounding factors, and (3) Exclusion restriction; the instrument is only associated with the outcome through the exposure^19^. We confirmed robust associations between the PRSs for both 25(OH)D and BMI in UKB, and BMI in HUNT. Limited measurements prevented us from testing the association with 25(OH)D in HUNT. In a previous study, a PRS based on three SNPs explained 3.7% of the variations of the 25(OH)D levels in a HUNT-subsample (n∼6300)^53^. Evaluating associations between PRSs and known confounders of the observational association are often used to partly assess the second assumption^19^. In our study, the BMI-PRS was weakly associated with 25(OH)D, age and sex, and the vitD-PRS weakly associated with BMI in UKB. However, the coefficients were small relative to the main effect on BMI. We can, however, not exclude associations with unknown or unmeasured confounders. Further, we recognize that the association between BMI-PRS and 25(OH)D, as well as the vitD-PRS and BMI, may have unknown effects on the interaction analysis.

When pleiotropy is present, the third assumption is violated^19^. Both instruments in our primary analyses have previously been used in one-sample MR with psoriasis as outcome and found to be robust to pleiotropy^11,37^. Instruments compiling variants in genes with well-known functional link to the exposure reduces risk of pleiotropy. Ideally, we would have included a such focused PRS for BMI as well. However, this was not attempted as most BMI associated variants have unknown functions^54,55^.

Factorial MR has low power to detect true interaction, even in large samples^20,21^. The difference in measured BMI (∼1.6 kg/m^2^) and 25(OH)D (∼7 nmol/L) between the groups in the 2×2 factorial MR analysis may be too small to detect interaction effects in our study. Comparing groups with genetically predicted larger differences than what is possible using common variants could provide more robust answers. Moreover, in simulation studies, an extension of two-stage least squares approach has been shown to be more efficient for analysing interactions^21^. However, such methods are not yet developed for binary outcomes^21^.

As discussed above, our findings reflect assumingly stable contribution of genetic risk over the life-course. This assumption may not hold, as the genetic contribution to BMI or 25(OH)D may differ with the state of the individual (e.g. with concurrent disease), at different time-points during development, or according to environmental exposures (like UV-availability). It is also plausible that the effect of overweight/vitamin D deficiency on psoriasis risk varies through life. Our data did not enable us to incorporate such potential time-varying effects. Life-course studies measuring BMI and 25(OH)D at several time-points may increase our understanding. Research on multiple sclerosis indicates crucial time-points for the effect of vitamin D on disease risk during childhood or adolescence^56^. To our knowledge, no such study on psoriasis risk has been performed.

Cohort studies tend to recruit the healthiest in the population^57,58^, and are thereby at risk for selection bias. However, we find it reasonable to assume that the potential effect of participation on BMI, 25(OH)D and genetic variants are independent of outcome status in our study. Any substantial impact on relative differences is therefore unlikely^59^. The consistency of our finding in both cohorts is also reassuring. By using a case definition combining self-report and different sources of diagnosis data, we aimed to include as many psoriasis cases as possible. This case definition has not been validated. Thus, the extent of outcome misclassification is unknown. It is, however, assumable that any expected outcome misclassification is non-differential (i.e. independent of both BMI, 25[OH]D and genetic variants). This may have biased the estimated ORs towards the null^41^.

The less clear trend between factorial groups in the 2×2 analysis in HUNT may be attributed to higher random error due to the smaller sample size. Additionally, the high degree of relatedness in the HUNT sample may result in over-precision and family-level bias in our estimate. However, we expect this to be minimal. Some variation between UKB and HUNT may also be due to differences in case definitions, as 55 % of the UKB sample lacked primary care data, and, that self-report in UKB only captures doctor diagnosed psoriasis. The study includes only white subjects of European ancestry and therefore may not be generalizable to other populations.

One strength of our study is that the use of genetic variants as proxies reduces the potential for confounding compared with traditional observational studies. Second, we utilize large-scale population-based cohorts compiling thousands of cases and controls, and thereby any large interaction effect is likely to be detected. We have shown consistent RERI estimates across two independent cohorts and between PRS-combinations. Our RERI estimates shows similar results in both 2×2 and continuous MR approaches, as well as in the observational analysis.

## Conclusions

The results from this factorial MR study suggest that there is no relative excess risk for psoriasis due to interaction between BMI and 25(OH)D. That is, the combined effect did not exceed the additive effect of the two factors. Interventions to reduce overweight and vitamin D deficiency in the population, may both influence psoriasis incidence, but the benefit of combined intervention cannot be expected to exceed the additive effect of the two. Considering the small differences in actual BMI and 25(OH)D between the factorial groups, small interaction effects or other threshold effects may have been undetected.

## Data Availability Statement

Data may be obtained from a third party and are not publicly available. The UK Biobank resource is available to bona fide researchers for health-related research in the public interest (https://www.ukbiobank.ac.uk/enable-your-research). Access to data from the HUNT Study can be given to researchers with a PhD associated with Norwegian research institutes upon application to HUNT’s Data Access Committee, given approval by a Regional Committee for Medical and Health Research Ethics. Researchers outside of Norway are welcome to apply for the use of HUNT data in cooperation with a Norwegian Principal Investigator (https://www.ntnu.edu/hunt/data).

## Conflict of Interest Statement

### Funding sources that supported the work

Marita Jenssen, Kjersti Danielsen and Anne-Sofie Furberg have received research grants from Northern Norway Regional Health Authority (grant number HNF1361-17 (MJ) and SFP1167-14 (KD, ASF), the Odd Berg Medical Research Foundation (MJ, KD) and the Norwegian Society of Dermatology and Venereology (MJ). Mari Løset and Ben Brumpton are supported by grants from the Liaison Committee for Education, Research and Innovation in Central Norway and the Joint Research Committee between St Olavs Hospital and the Faculty of Medicine and Health Sciences, NTNU. Yi-Qian Sun was supported by a Researcher grant from the Liaison Committee for Education, Research, and Innovation in Central Norway (project ID 2018/42794). The other authors state no funding supporting the work reported in the manuscript.

The funders were not involved in the design and conduct of the study; nor in the collection, management, analysis, and interpretation of the data; nor in the preparation, review, or approval of the manuscript; nor in the decision to submit the manuscript for publication.

## Acknowledgements

We want to thank the participants and staff of the UK Biobank and the Trøndelag Health Study.

The Trøndelag Health Study (HUNT) is a collaboration between HUNT Research Center (Faculty of Medicine and Health Sciences, NTNU, Norwegian University of Science and Technology), Trøndelag County Council, Central Norway Regional Health Authority, and the Norwegian Institute of Public Health. The genotyping in HUNT was financed by the National Institutes of Health; University of Michigan; the Research Council of Norway; the Liaison Committee for Education, Research and Innovation in Central Norway; and the Joint Research Committee between St Olavs hospital and the Faculty of Medicine and Health Sciences, NTNU. Analyses were performed in digital labs at HUNT Cloud, NTNU, Norwegian University of Science and Technology, Trondheim, Norway.

## CredIT statement

Conceptualization: MJ, ML, BOÅ, KD, BMB; Methodology: NA, BMB; Formal analysis: MJ, NA, BMB; Data Curation: LT, BMB; Writing - Original Draft: MJ; Writing - Review & Editing: NA, ML, BOÅ, LT, OJGB, XMM, YQS, ASF, RJ, TW, KD, BMB; Visualization: MJ, NA, BMB; Supervision: KD, BMB; Funding acquisition: KD, BMB

## Supplementary material

Online supplementary material.

