## Supplementary material for "Exploring interaction between genetically predicted body mass index and serum 25-hydroxyvitamin D levels on the odds for psoriasis in UK Biobank and the HUNT Study: A factorial Mendelian randomisation study": Online supplementary

**Online supplementary material**

Content

### Flow-charts

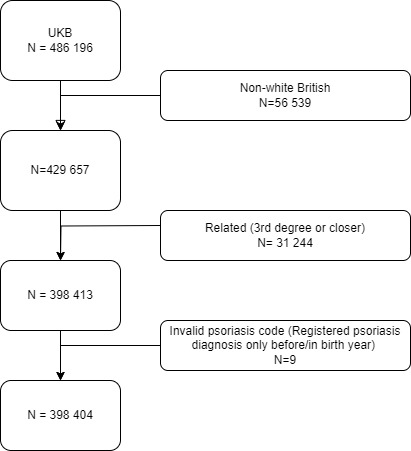

Figure S1. Flow-chart of the study sample selection. The UK Biobank (UKB).

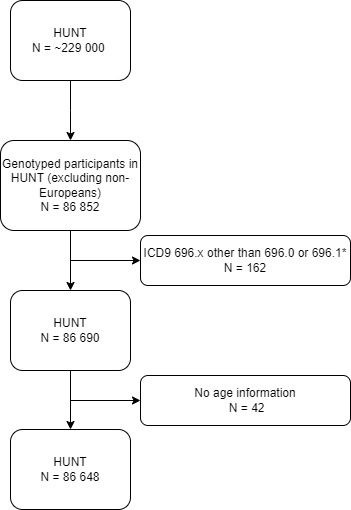

Figure S2. Flow-chart of the study sample selection. The Trøndelag Health study (HUNT).

*participants with ICD9 codes 696.2 Parapsoriasis, 696.3 Pityriasis rosea, 696.4 Pityriasis rubra pilaris, 696.5 Other and unspecified pityriasis, or 696.8 Other psoriasis and similar disorders were excluded.

### UK Biobank (UKB) field IDs used in the study

| **UKB data-field** | **Field-ID** |
| --- | --- |
| Ethnicity (White British) | 21000 |
| Sex | 31 |
| Date of first L40 reported (psoriasis) | 131742 |
| Source of report of L40 | 131743 |
| Age at assessment | 21003 |
| Body mass index (BMI) | 21001 |
| Serum 25-hydroxyvitamin D | 30890 |
| Month of attending assessment centre | 55 |
| Genotype measurement batch | 22000 |
| Genetic principal components | 22009 |

### Psoriasis classification in UKB

| Table S1  Source of psoriasis diagnosis UK Biobank | | |
| --- | --- | --- |
|  | **N** | **%** |
| Primary care only | 4 573 | 37.5 |
| Primary care and other source(s) | 863 | 7.1 |
| Hospital admissions data only | 1730 | 14.2 |
| Hospital admissions data and other source(s) | 64 | 0.5 |
| Self-report only | 2 559 | 21.0 |
| Self-report and other source(s) | 2 418 | 19.8 |

### Psoriasis classification in The Trøndelag Health study (HUNT)

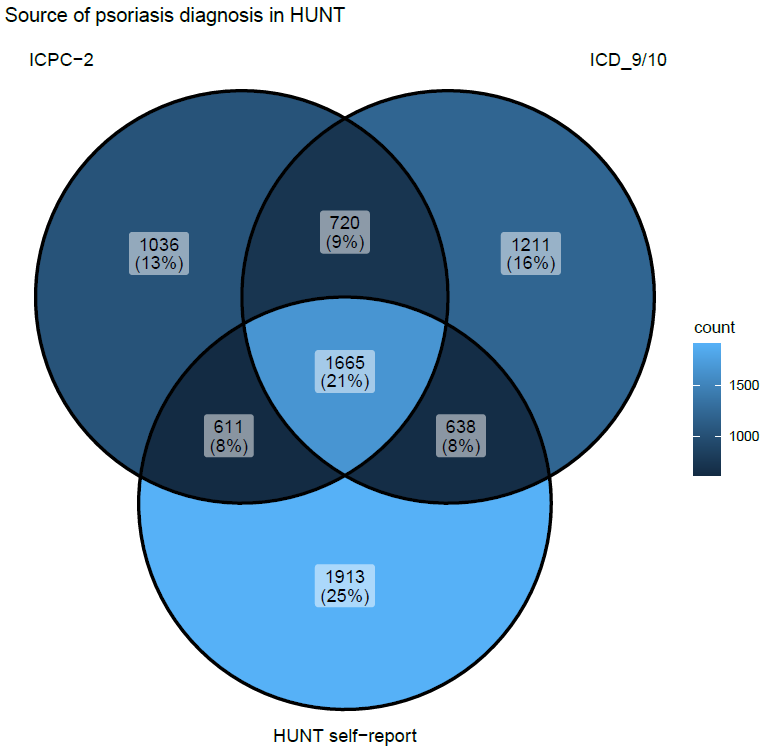

Figure S3. Venn-diagram showing the source of psoriasis diagnosis in HUNT. Numbers shown as count (%).

ICPC-2 = International Classification of Primary Care, 2nd Edition. ICD_9/10 = International Classification of Diseases, Ninth and Tenth Revision

### Additional details regarding the genetic instruments

We calculated polygenic risk scores (PRSs) for body mass index (BMI) and 25-hydroxyvitamin D (25[OH]D) for each participant as the sum of trait increasing alleles, weighted by the genome wide association study (GWAS)-effect size of the respective single nucleotide polymorphism (SNP)(Burgess and Thompson, 2013).

For our primary analysis, we developed a PRS for BMI (hereafter referred to as BMI-PRS) combining 941 SNPs robustly associated with BMI in participants of European decent in the most recent GWAS meta-analysis (also including UKB participants)(Yengo et al., 2018). Of these, 940 variants were available in UKB and all in HUNT. In sensitivity analysis, we developed another BMI-PRS combining 77 SNPs robustly associated with BMI in the most recent GWAS which did not include UKB-participants(Locke et al., 2015), and thereby avoiding internal weights(Burgess and Thompson, 2013). Of these, all were available in UKB, and 76 were available in HUNT. Both of the BMI-PRSs has previously been used in a one-sample mendelian randomisation (MR) study in HUNT and UKB, confirming a causal association between BMI and psoriasis, also when applying pleiotropy robust methods(Budu-Aggrey et al., 2019).

For the PRS for 25(OH)D (hereafter referred to as vitD-PRS) we presented combinations of SNPs used in previous MR studies, where researchers have used different approaches in the selection of SNPs. In our primary analysis the vitD-PRS included 21 SNPs from genes with well-known functions in the vitamin D metabolism (*CYP2R1*, *DHCD7*, *CYP24A1* and *GC*), selected by Sofianopoulou et al. using a step-wise procedure based on genome-wide significant association (p <5 x 10^-8^) with 25(OH)D(Sofianopoulou et al., 2021). By selecting variants in genes with well-known functions, risk of pleiotropy is minimized(Sofianopoulou et al., 2021). In sensitivity analyses, we used a vitD-PRS including 35 SNPs, selected in previous MR studies by Zhou and Hyppönen(Zhou and Hyppönen, 2023, Zhou et al., 2022). They evaluated genome-wide significant SNPs associated with 25(OH)D in a recent GWAS(Revez et al., 2020), and selected those in which the association were replicated in an earlier GWAS(Jiang et al., 2018). Thus, they ensured the robustness in the GWAS signal in an external data source, and avoided the use of internal weights(Burgess and Thompson, 2013, Zhou and Hyppönen, 2023). We also explored the impact of using a more extensive vitD-PRS including 71 genome-wide significant SNPs from a recent GWAS(Manousaki et al., 2020), which were explored in Sofianopoulou’s study(Sofianopoulou et al., 2021). All selected variants were available in UKB. In HUNT, 19 of 21, 32 of 35, and 61 of 71 variants were available, respectively.

The SNPs and weights used in our analyses can be found in the supplementary Excel-file.

### Association between PRSs and possible confounders of the observational association

***BMI-PRS***

In UKB the BMI-PRS (941 SNPs) was weakly associated with 25(OH)D, and even more weakly with age and sex (Table S2a).

In HUNT, the BMI-PRS (941 SNPs) was associated with age, but with sex (Table S2a).

| Table S2a  Association between the polygenic risk score for BMI (BMI-PRS)  and possible confounders of the observational association in UKB and HUNT. | | | | |
| --- | --- | --- | --- | --- |
|  | UKB | | HUNT | |
| **Dependent variable^1^** | **Beta coefficient per 1 SD unit increase in BMI-PRS^1^** | **95 % CI** | **Beta coefficient per 1 SD unit increase in BMI-PRS^1^** | **95 % CI** |
| 25(OH)D | -0.03 | -0.03, -0.02 | NA | NA |
| Age | -0.002 | -0.004, -0.001 | -0.45 | -0.76, -0.13 |
| Sex | 0.0001 | 0.00002, 0.0002 | 0.02 | 0.01, 0.02 |
| ^1^ Beta coefficients from separate univariable linear models (ANOVA) for each of the listed dependent variables.  BMI-PRS= polygenic risk score for body mass index. UKB= UK Biobank. HUNT = the Trøndelag Health Study. 25(OH)D = 25-hydroxyvitamin D. SD = standard deviation. CI = confidence interval. NA=not applicable. | | | | |

***VitD-PRS***

In UKB, the vitD-PRS (21 SNPs) was weakly associated with BMI (Table S2b). It was not associated with age or sex (Table S2b).

In HUNT, the vitD-PRS (21 SNPs) not associated with BMI, nor with age or sex (Table S2b).

| Table S2b  Association between the polygenic risk score for 25-hydroxyvitamin D (vitD-PRS)  and possible confounders of the observational association in UKB and HUNT. | | | | |
| --- | --- | --- | --- | --- |
|  | UKB | | HUNT | |
| **Dependent variable^1^** | **Beta coefficient per 1 SD unit increase in vitD-PRS^1^** | **95 % CI** | **Beta coefficient per 1 SD unit increase in vitD-PRS^1^** | **95 % CI** |
| BMI | 0.02 | 0.006, 0.03 | -0.06 | -0.19, 0.07 |
| Age | -0.001 | -0.02, 0.02 | -0.007 | -0.51, 0.53 |
| Sex | 0.001 | -0.0002, 0.002 | -0.01 | -0.03, 0.003 |
| ^1^ Beta coefficients from separate univariable linear models (ANOVA) for each of the listed dependent variables.  vitD-PRS=polygenic risk score for 25-hydroxyvitamin D. UKB=UK Biobank. HUNT= the Trøndelag Health Study. BMI=body mass index. SD=standard deviation. CI=confidence interval. | | | | |

### Sensitivity analyses

Results from sensitivity analyses comparing results from factorial MR analyses using different combinations of PRSs for BMI and 25(OH)D in UKB (Table S3a and S4a) and HUNT (Table S3b and S4b). As a measure of additive interaction, we present estimated values for relative excess risk due to interaction (RERI) across the combinations. RERI estimates are valid when all estimated ORs are above 1(Knol et al., 2011). In the 2x2 MR analysis in HUNT, some of the combination has an estimated OR below 1 for the Factorial group 1 in combinations including the focused (19 SNPs) and the extended (61 SNPs) vitD-PRS. These RERI estimates may not be valid, and must be interpreted with caution. We therefore refrain from trusting the borderline significant RERI estimate for the 941-61 combination in HUNT (Table S3b).

| Table S3a  2x2 factorial mendelian randomisation analysis comparing odds ratio estimates for psoriasis using different sets of polygenic risk scores for body mass index and 25-hydroxyvitamin D. The UK Biobank. | | | | | | |
| --- | --- | --- | --- | --- | --- | --- |
| BMI-PRS^1^ | 77 | | | 940 | | |
| VitD-PRS^1^ | 21 | 35 | 71 | 21 | 35 | 71 |
|  | **OR (95% CI)** | **OR (95% CI)** | **OR (95% CI)** | **OR (95% CI)** | **OR (95% CI)** | **OR (95% CI)** |
| Factorial group 0 | 1 (ref.) | 1 (ref.) | 1 (ref.) | 1 (ref.) | 1 (ref.) | 1 (ref.) |
| Factorial group 1 | 1.05 (1.00, 1.10) | 1.04 (0.99, 1.09) | 1.05 (1.00, 1.11) | 1.04 (0.99, 1.09) | 1.07 (1.02, 1.13) | 1.03 (0.98, 1.09) |
| Factorial group 2 | 1.03 (0.98, 1.09) | 1.02 (0.97, 1.08) | 1.05 (0.99, 1.10) | 1.07 (1.02, 1.13) | 1.10 (1.05, 1.16) | 1.08 (1.02, 1.13) |
| Factorial group 3 | 1.07 (1.02, 1.13) | 1.08 (1.02, 1.13) | 1.06 (1.01, 1.12) | 1.12 (1.07, 1.18) | 1.13 (1.07, 1.19) | 1.11 (1.06, 1.17) |
|  | **RERI (95% CI)** | **RERI (95% CI)** | **RERI (95% CI)** | **RERI (95% CI)** | **RERI (95% CI)** | **RERI (95% CI)** |
|  | -0.01 (-0.08, 0.07) | 0.02 (-0.06, 0.09) | -0.03 (-0.11, 0.04) | 0.01 (-0.06, 0.09) | -0.05 (-0.13, 0.03) | 0.01 (-0.07, 0.08) |
| ^1^ Digits refers to the number of single nucleotide polymorphisms (SNPs) included in the polygenic risk scores (PRSs). For the BMI-PRSs 77 of 77 and 940 of 941 selected SNPs were available, for the VitD-PRSs all selected variants were available. All PRSs were dichotomized at the median value, and combined to four factorial groups:  Factorial group 0 = PRS-BMI below median, PRS-vitD above median; expected to have the lowest risk for psoriasis, and therefore chosen as reference group.  Factorial group 1 = PRS-BMI below median, PRS-vitD below median.  Factorial group 2 = PRS-BMI above median, PRS-vitD above median.  Factorial group 3 = PRS-BMI above median, PRS-vitD below median.  All models adjusted for age, sex, genetic batch and principal components 1 to 20.  BMI-PRS=polygenic risk score for body mass index. vitD-PRS= polygenic risk score for 25-hydroxyvitamin D. OR=odds ratio. CI=confidence interval. Ref.=reference.  RERI=relative excess risk due to interaction. Interaction exists if RERI ≠ 0. | | | | | | |

| Table S3b  2x2 factorial mendelian randomisation analysis comparing odds ratio estimates for psoriasis using different sets of polygenic risk scores for body mass index and 25-hydroxyvitamin D. The HUNT Study. | | | | | | |
| --- | --- | --- | --- | --- | --- | --- |
| BMI-PRS^1^ | 76 | | | 941 | | |
| VitD-PRS^1^ | 19 | 32 | 61 | 19 | 32 | 61 |
|  | **OR (95% CI)** | **OR (95% CI)** | **OR (95% CI)** | **OR (95% CI)** | **OR (95% CI)** | **OR (95% CI)** |
| Factorial group 0 | 1 (ref.) | 1 (ref.) | 1 (ref.) | 1 (ref.) | 1 (ref.) | 1 (ref.) |
| Factorial group 1 | 0.96 (0.90, 1.02) | 1.02 (0.95, 1.09) | 0.98 (0.91, 1.04) | 0.99 (0.93, 1.06) | 1.00 (0.93, 1.07) | 0.95 (0.89, 1.01) |
| Factorial group 2 | 1.07 (1.00, 1.14) | 1.09 (1.02, 1.17) | 1.06 (0.99, 1.13) | 1.09 (1.02, 1.17) | 1.06 (0.99, 1.13) | 1.02 (0.95, 1.09) |
| Factorial group 3 | 1.06 (0.99, 1.13) | 1.10 (1.03, 1.17) | 1.08 (1.02, 1.16) | 1.04 (0.97, 1.11) | 1.08 (1.01, 1.16) | 1.07 (1.00, 1.14) |
|  | **RERI (95% CI)** | **RERI (95% CI)** | **RERI (95% CI)** | **RERI (95% CI)** | **RERI (95% CI)** | **RERI (95% CI)** |
|  | 0.03 (-0.07, 0.12) | -0.01 (-0.11, 0.09) | 0.05 (-0.05, 0.14) | -0.04 (-0.14, 0.06) | 0.03 (-0.07, 0.12) | 0.10 (0.01, 0.19) |
| ^1^ Digits refers to the number of single nucleotide polymorphisms (SNPs) included in the respective polygenic risk score (PRS). For the BMI-PRSs, 76 of 77 and 941 of 941 selected SNPs were available, for the VitD-PRSs 19 of 21, 32 of 35, and 61 of 71 selected SNPs were available. All PRSs were dichotomized at the median value, and combined to four factorial groups:  Factorial group 0 = PRS-BMI below median, PRS-vitD above median; expected to have the lowest risk for psoriasis, and therefore chosen as reference group.  Factorial group 1 = PRS-BMI below median, PRS-vitD below median.  Factorial group 2 = PRS-BMI above median, PRS-vitD above median.  Factorial group 3 = PRS-BMI above median, PRS-vitD below median.  All models adjusted for age, sex, genetic batch and principal components 1 to 20.  HUNT=The Trøndelag Health Study (HUNT2-4). BMI-PRS=polygenic risk score for body mass index. VitD-PRS = polygenic score for 25-hydroxyvitamin D. OR=odds ratio. CI=confidence interval.  RERI=relative excess risk due to interaction. Interaction exists if RERI ≠ 0. | | | | | | |

| Table S4a  Continuous factorial mendelian randomisation analyses comparing odds ratio estimates for psoriasis using different sets of polygenic risk scores for body mass index and 25-hydroxyvitamin D, included in the model as standardised continuous variables and their cross-product term. The UK Biobank. | | | | | | | |
| --- | --- | --- | --- | --- | --- | --- | --- |
| BMI-PRS^1^ | 77 | | | | 940 | | |
| VitD-PRS^1^ | 21 | 35 | 71 | 21 | | 35 | 71 |
|  | **OR (95% CI)** | **OR (95% CI)** | **OR (95% CI)** | **OR (95% CI)** | | **OR (95% CI)** | **OR (95% CI)** |
| BMI-PRS (per 1 SD increase) | 1.03 (1.01, 1.05) | 1.03 (1.01, 1.05) | 1.03 (1.01, 1.05) | 1.04 (1.02, 1.06) | | 1.04 (1.02, 1.06) | 1.04 (1.02, 1.06) |
| VitD-PRS (per 1 SD decrease) | 1.02 (1.00, 1.04) | 1.02 (1.00, 1.04) | 1.02 (1.00, 1.04) | 1.02 (1.00, 1.04) | | 1.02 (1.00, 1.04) | 1.02 (1.00, 1.04) |
| Cross-product  (BMI-PRS*VitD-PRS) | 1.00 (0.99, 1.02) | 1.00 (0.98, 1.02) | 1.00 (0.98, 1.02) | 1.00 (0.98, 1.01) | | 0.99 (0.97, 1.01) | 1.00 (0.98, 1.02) |
|  | **OR** | **OR** | **OR** | **OR** | | **OR** | **OR** |
| Additive term  (BMI-PRS+VitD-PRS+cross product^2^) | 1.06 | 1.05 | 1.05 | 1.06 | | 1.05 | 1.06 |
|  | **RERI (95% CI)** | **RERI (95% CI)** | **RERI (95% CI)** | **RERI (95% CI)** | | **RERI (95% CI)** | **RERI (95% CI)** |
|  | 0.00 (-0.01, 0.02) | 0.00 (-0.02, 0.02) | 0.00 (-0.02, 0.02) | 0.00 (-0.02, 0.02) | | -0.01 (-0.03, 0.01) | 0.00 (-0.02, 0.02) |
| ^1^ Digits refers to the number of single nucleotide polymorphisms (SNPs) included in the respective polygenic risk scores (PRSs). For the BMI-PRSs, 77 of 77 and 940 of 941 selected SNPs were available, for the VitD-PRSs all selected variants were available.  ^2^ Calculated by exponentiating the sum of estimated beta-values for BMI-PRS, VitD-PRS and their cross-product (exp[b1+b2+b3]).  All models adjusted for age, sex, genetic batch and principal components 1 to 20.  BMI-PRS=polygenic risk score for body mass index. vitD-PRS= polygenic risk score for 25-hydroxyvitamin D. OR=odds ratio. CI=confidence interval. SD=standard deviation. RERI=relative excess risk due to interaction. Interaction exists if RERI ≠ 0. | | | | | | | |

| Table S4b Continuous factorial mendelian randomisation analyses comparing odds ratio estimates for psoriasis using different sets of polygenic risk scores for body mass index and 25-hydroxyvitamin D, included in the model as standardised continuous variables and their cross-product term. The HUNT Study. | | | | | | |
| --- | --- | --- | --- | --- | --- | --- |
| BMI-PRS^1^ | 76 | | | 941 | | |
| VitD-PRS^1^ | 19 | 32 | 61 | 19 | 32 | 61 |
|  | **OR (95% CI)** | **OR (95% CI)** | **OR (95% CI)** | **OR (95% CI)** | **OR (95% CI)** | **OR (95% CI)** |
| BMI-PRS (per 1 SD increase) | 1.05 (1.02, 1.07) | 1.05 (1.02, 1.07) | 1.05 (1.02, 1.07) | 1.04 (1.01, 1.06) | 1.04 (1.01, 1.06) | 1.04 (1.01, 1.06) |
| VitD-PRS (per 1 SD decrease) | 1.02 (1.00, 1.05) | 0.99 (0.97, 1.02) | 0.98 (0.96, 1.00) | 1.02 (1.00, 1.05) | 0.99 (0.97, 1.02) | 0.98 (0.96, 1.00) |
| Cross-product (BMI-PRS*VitD-PRS) | 1.00 (0.97, 1.02) | 1.00 (0.98, 1.03) | 1.00 (0.97, 1.02) | 1.00 (0.97, 1.02) | 1.00 (0.97, 1.02) | 1.00 (0.97, 1.02) |
|  | **OR** | **OR** | **OR** | **OR** | **OR** | **OR** |
| Additive term  (BMI-PRS+VitD-PRS+cross product^2^) | 1.07 | 1.04 | 1.02 | 1.06 | 1.03 | 1.01 |
|  | **RERI (95% CI)** | **RERI (95% CI)** | **RERI (95% CI)** | **RERI (95% CI)** | **RERI (95% CI)** | **RERI (95% CI)** |
|  | -0.00 (-0.03, 0.02) | 0.00 (-0.02, 0.03) | -0.00 (-0.03, 0.02) | -0.00 (-0.03, 0.02) | -0.01 (-0.03, 0.02) | -0.00 (-0.03, 0.02) |
| ^1^ Digits refers to the number of single nucleotide polymorphisms (SNPs) included in the polygenic risk scores (PRS). For the BMI-PRSs 76 of 77 and 941 of 941 selected SNPs were available, for the VitD-PRSs 19 of 21, 32 of 35, and 61 of 71 selected SNPs were available.  ^2^ Calculated by exponentiating the sum of estimated beta-values for BMI-PRS, VitD-PRS and their cross-product (exp[b1+b2+b3]).  All models adjusted for age, sex, genetic batch and principal components 1 to 20.  HUNT= The Trøndelag Health Study (HUNT2-4). BMI-PRS=polygenic risk score for body mass index. vitD-PRS= polygenic risk score for 25-hydroxyvitamin D. OR=odds ratio. CI=confidence interval. SD=standard deviation. RERI=relative excess risk due to interaction. Interaction exists if RERI ≠ 0. | | | | | | |

### Observational data from UKB

In UKB, a 10 nmol/L decrease in measured serum 25(OH)D was associated with 3% higher odds for psoriasis (OR 1.03, 95% confidence interval [CI] 1.02, 1.04), and a 5 kg/m^2^ increase in measured BMI was associated with 19% higher odds for psoriasis (OR 1.19, 95% CI 1.17, 1.22) (Table S5a). The combined effect of the two factors summed to 22% higher odds for psoriasis (OR 1.22, 95% CI 1.09, 1.37), supporting exact additivity in the observational association (RERI 0.00, 95% CI -0.01, 0.03). The same was found in 2x2 analysis (Table S5b). The single effect of having serum 25(OH)D <25 nmol/L was associated with 21% higher odds for psoriasis (OR 1.21, 95% CI 1.09, 1.33) and the single effect of BMI>27.5 kg/m^2^ was associated with 38% higher odds for psoriasis (OR 1.38, 95% CI 1.33, 1.43). Whilst having both 25(OH)D <25 nmol/L and BMI>27.5 was associated with 59% higher odds for psoriasis (OR 1.59, 95% CI, 1.40, 1.80) compared with having both 25(OH)D≥25 nmol/L and BMI ≤27.5 kg/m^2^ (RERI 0.05, 95% CI -0.21, 0.32).

| Table S5a  Odds ratios for psoriasis per 10 nmol/L decrease in measured serum 25-hydroxyvitamin D and 5 kg/m2 increase in measured body mass index. Cross-sectional analysis of data from the UK Biobank (n=12 207 psoriasis/386 197 non-psoriasis*). | | |
| --- | --- | --- |
| **Predictor** | **OR (95% CI)^1^** | **OR (95% CI) ^1^** |
| 25(OH)D, per 10 nmol/L decrease | 1.03 (1.02, 1.04) | 1.03 (0.98, 1.09) |
| BMI, per 5 kg/m2 increase | 1.19 (1.17, 1.22) | 1.18 (1.13, 1.25) |
| BMI*25(OH)D (cross-product) |  | 1.00 (0.99, 1.01) |
| Additive interaction between  25(OH)D and BMI^2^ |  | 1.22 (1.09, 1.37) |
|  |  | **RERI (95% CI)** |
|  |  | 0.00 (-0.01, 0.03) |
| ^1^ Logistic regression model.  Adjusted for age and sex.  ^2^ Calculated by exponentiating the sum of estimated beta-values for 25(OH)D, BMI and their cross-product (exp[b1+b2+b3]).  25(OH)D=25-hydroxyvitamin D (nmol/L). BMI=body mass index (kg/m^2^). OR=odds ratio. CI=confidence interval. RERI=relative excess risk due to interaction. Interaction exists if RERI ≠ 0.  *counts differ from the Mendelian randomisation analyses because of missing measured values (BMI n=1262; 25(OH)D n=35247) | | |

| Table S5b  Odds ratios for psoriasis in groups based on dichotomised measured body mass index and 25-hydroxyvitamin D levels.  Cross-sectional analysis of observational data from The UK Biobank (n=12 207 psoriasis/386 197 non-psoriasis*). | | | | | | |
| --- | --- | --- | --- | --- | --- | --- |
|  | **BMI >27.5** | **25(OH)D <25^1^** | **Psoriasis** | **Non-psoriasis** | **OR (95% CI)**  (Unadjusted) | **OR (95% CI)**  (Adjusted^1^) |
| Single associations | - | No | 10669 | 339125 | 1 (Reference) | 1 (Reference) |
|  | - | Yes | 435 | 11666 | 1.19 (1.07, 1.31) | 1.21 (1.09, 1.33) |
|  | No | - | 5449 | 220938 | 1 (Reference) | 1 (Reference) |
|  | Yes | - | 5655 | 165259 | 1.40 (1.35, 1.46) | 1.38 (1.33, 1.43) |
| Combined associations | No | No | 5287 | 196501 | 1 (Reference) | 1 (Reference) |
|  | No | Yes | 162 | 5255 | 1.14 (0.98, 1.34) | 1.16 (0.99, 1.35) |
|  | Yes | No | 5382 | 142624 | 1.40 (1.35, 1.46) | 1.38 (1.33, 1.43) |
|  | Yes | Yes | 273 | 6411 | 1.58 (1.40, 1.79) | 1.59 (1.40, 1.80) |
|  |  |  |  |  | **RERI (95% CI)** | **RERI (95% CI)** |
|  |  |  |  |  | 0.03 (-0.23, 0.30) | 0.05 (-0.21, 0.32) |
| ^1^ Season adjusted 25(OH)D levels  ^2^ Adjusted for age and sex.  BMI=body mass index (kg/m^2^). 25(OH)D=25-hydroxyvitamin D (nmol/L). OR=odds ratio. CI=confidence interval.  RERI=relative excess risk due to interaction. Interaction exists if RERI ≠ 0.  *counts differ from factorial Mendelian randomisation analyses because of missing measured values (BMI n=1262; 25(OH)D n=35247) | | | | | | |
